# Prenatal Exposure to Gestational Diabetes Mellitus is Associated with Greater Pre-pubertal BMI Growth and Faster Post-pubertal Cortical Thinning During Peri-adolescence

**DOI:** 10.1101/2025.03.25.25324581

**Authors:** Eustace Hsu, Trevor A. Pickering, Shan Luo

## Abstract

**Background:** The longitudinal trajectory of body mass index (BMI) and brain structure development during peri-adolescence is not clearly defined in offspring prenatally exposed to gestational diabetes mellitus (GDM) vs. un-exposed offspring.

**Methods:** Participants between age 9 and 10 years (N=9,583) were included from the Adolescent Brain and Cognitive Development (ABCD) Study and followed yearly though 4-year follow-up. GDM and puberty status were self-reported. BMI was calculated yearly, and MRI assessed brain structure biennially. Mixed-effects models analyzed trajectories of BMI and brain structural measures between groups controlling for sociodemographic covariates, and linear spline was defined with a knot at onset of puberty.

**Results:** There was an interaction of exposure by age in change in BMI [β (95% CI) = 0.032 (0.008, 0.056), *P*=0.009] and mean cortical thickness [β (95% CI) = −0.038 (−0.071, −0.004), *P*=0.027]. The former was driven by greater pre-pubertal increases in BMI [β (95% CI) = 0.051 (0.002, 0.100), *P*=0.043], whereas the latter was driven by faster post-pubertal declines in cortical thickness among GDM-exposed offspring [β (95% CI) = −0.051 (−0.095, −0.007), *P*=0.046].

**Conclusion:** Prenatal GDM exposure is associated with greater pre-pubertal increases in BMI and faster post-pubertal cortical thinning in youth age between 9 and 15.

**Practitioner Points:**

– Prenatal GDM exposure is associated with greater pre-pubertal increases in BMI and faster post-pubertal cortical thinning in youth age between 9 and 15.
– It is important to recognize puberty as a window of vulnerability for altered brain development among youth prenatally exposed to GDM.

## Introduction

Prenatal exposure to gestational diabetes mellitus (GDM) is an established risk factor for adolescent obesity (1–9). Observational studies have reported larger measures of adiposity markers in GDM-exposed offspring (vs. un-exposed), but these studies have predominantly been cross-sectional (6,7,10–12). Longitudinal studies have sought to identify where divergence in BMI growth trajectory between GDM-exposed and un-exposed offspring occurs. While comparisons of adiposity markers between groups during early childhood between birth and 5 years have not yielded clear results (13), longitudinal studies spanning across later childhood to adolescence demonstrate greater increases in BMI (10,14) and greater prevalence of overweight and obesity (15) among youth exposed to GDM compared with un-exposed youth (13). In particular, one longitudinal study identified the transition from childhood to adolescence (10-13 years) as the time window when GDM exerts the strongest effect upon BMI growth (16,17). Transition to adolescence or puberty is marked by significant changes in BMI growth. Considering this evidence, we hypothesize that onset of puberty represents a divergence point for BMI growth trajectory between GDM-exposed and un-exposed youth, which we will formally test in this study.

Transition to adolescence is also characterized by significant changes in brain development. Cortical and subcortical grey matter volume (GMV), cortical thickness (CT) and surface area (SA) peak ranging between early childhood and early adolescence and decrease throughout adolescence (18). And pubertal development is associated with brain structural development during the peri-adolescence period (18–20). However, it is currently unknown how prenatal exposure to GDM is related to longitudinal trajectory of brain structural development during peri-adolescence and the role of puberty in the impact of GDM exposure on brain structural development.

In this study, we leveraged longitudinal BMI and brain structural data from the Adolescent Brain and Cognitive Development Study® to examine the trajectory of BMI growth and brain structure development in GDM-exposed and un-exposed youth between the ages of 9 and 15 years. We hypothesize that prenatal exposure to GDM will be associated with greater longitudinal increases in BMI and greater decreases in structural measures such as GMV, CT and SA, and onset of puberty will be a divergence point between groups in the trajectory of BMI and brain structure measures.

## Methods

### Participants

The ABCD® Study is an ongoing longitudinal study of adolescent development, with data collection at 21 study sites across the U.S. Data for this project was obtained from the ABCD 5.1 release, which includes data from baseline, one-, two-, three- and four-year follow-up, where participants were 9-10 years old at baseline and as old as 15 years, 9 months at 4-year follow-up. Details of study design, recruitment, and inclusion/exclusion for the ABCD study have previously been reported (20,21).

Participants were excluded if they met any of the following criteria: not fluent in English, history of seizures, birth more than 12 weeks premature, birth weight less than 1200 grams, complications at birth, substance use disorder, intellectual disability, traumatic brain injury, lead poisoning, schizophrenia, major neurological disorders including cerebral palsy and muscular dystrophy, and other medical conditions considered exclusionary, beginning at reported onset (details see Supporting Figure 1).

**Figure 1.**
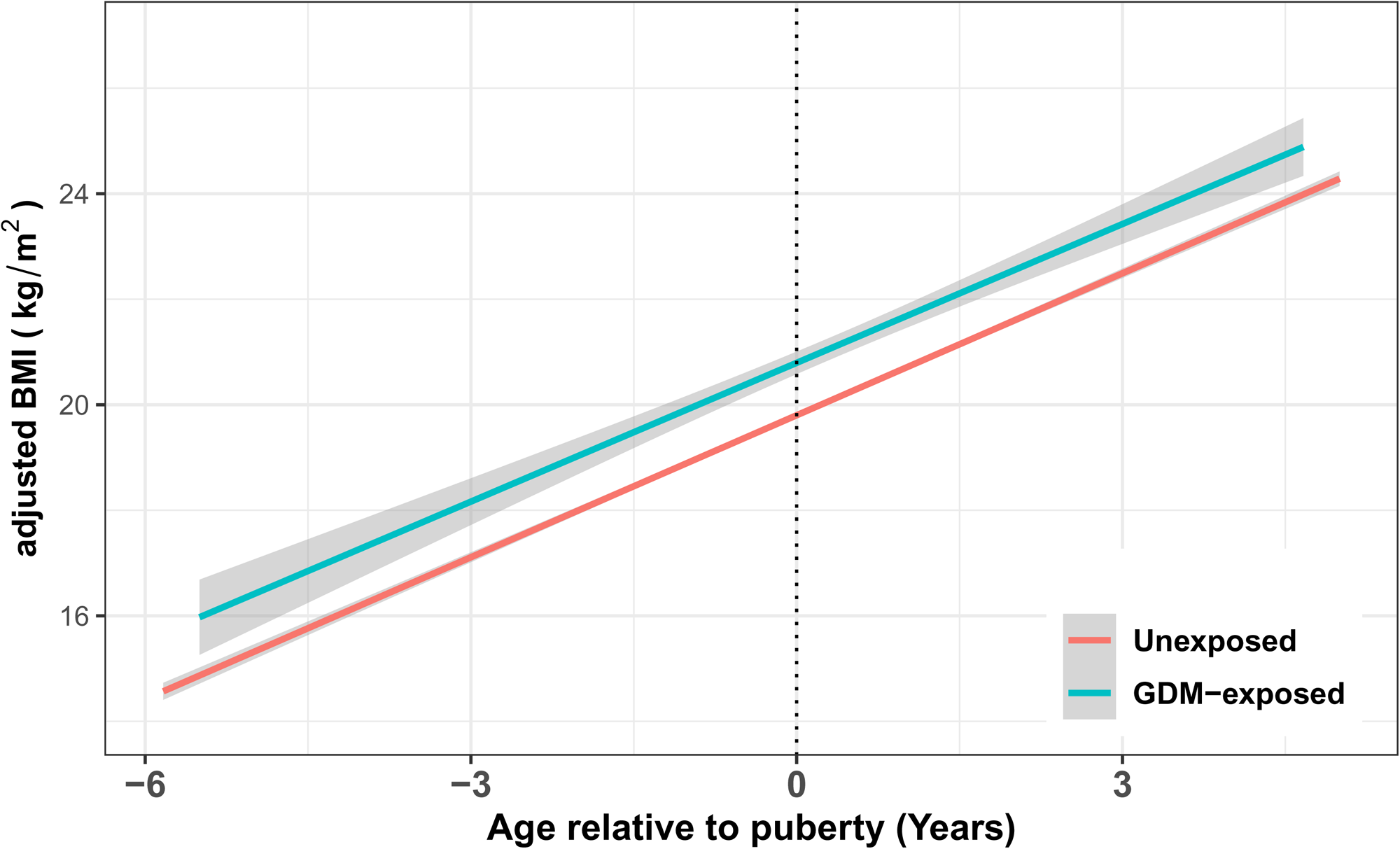
Relationship of maturation age (age relative to onset of puberty, x-axis) by gestational diabetes mellitus (GDM) exposure status interaction in body mass index (BMI) with adjustment for family ID and site ID as random effects, and sex, race, ethnicity, current pubertal status, current family income, and current parental education as fixed effects. Lines represent mixed-effects regression linear fit and shaded region represents one standard error (SE) region of regression fit.

### Gestational diabetes mellitus exposure

GDM exposure was self-reported by caregiver at initial baseline interview based on the question: “During the pregnancy with this child, did you/biological mother have pregnancy-related diabetes?”, and coded as a binary variable. And ∼88.8% of the data was reported by the biological mother.

### Pubertal Stage

Pubertal stage was reported by caregiver at baseline and yearly follow-up based on the pubertal development scale (PDS), an assessment which has been validated against the traditional tanner staging (23,24). The PDS consists of 5 questions regarding changes in height, body hair, skin, breast development, and menarche (female) or changes in height, body hair, skin, voice, and facial hair (male). PDS summary scores were calculated for girls and boys based on a subset of features (body hair and breast development among females, and body hair, voice, and facial hair among males), and subsequently converted to a Tanner Staging category score.

Individual onset of puberty was defined as the first follow-up with Tanner Staging ≥2. If pubertal stage was Tanner Stage ≥2 at baseline, then baseline age was used as onset of puberty. Maturation age was calculated as the age relative to age of onset of puberty. For participants with Tanner Stage = 1 at all recorded follow-ups, onset of puberty was imputed as 12 months or one year older than the age of the latest recorded follow-up, in order to preserve their inclusion in the study.

### Anthropometrics

Weight and height were recorded at yearly visits by trained staff, as the mean of two to three measurements. Measurements were converted to metric scale and used to calculate BMI (kg/m^2^). Modified BMI *z*-scores (BMI relative to CDC growth chart BMI median by age and sex), were calculated (25–27), and modified BMI *z*-score cut offs of < −4 and > 8 were excluded as biologically implausible values at each time point (27,28).

To identify longitudinal implausible measurements, sex-stratified, mixed effects linear spline models were conducted, with BMI modeled as a function of age, with race and ethnicity as fixed effects covariates, and subject as a random effect. A spline for age was modeled with a knot at 10 years, 6 months for females and 11 years, 8 months for males, to reflect the mean age of onset of puberty across the study. Studentized residuals were obtained from the model, and studentized residuals >|6| were identified as longitudinal implausible measurements and excluded [for all details see Boone-Heinonen 2019(27)].

### Neuroimaging

T1-weighted data was collected at baseline, 2-years, and 4-years follow-up using 3-Tesla scanners optimized and harmonized across multiple platforms (30). Cortical surface reconstruction and subcortical segmentation were completed at the ABCD® Data Analytics and Informatics Center (DAIC) via FreeSurfer (version **7.1.1**), producing estimates of interests for this study, including CT (mm), cortical SA (mm^2^), and cortical and subcortical GMV, and intracranial volume (ICV) (mm^3^). Quality control (QC) was conducted to evaluate cortical surface reconstruction for five categories of inaccuracy: severity of motion, intensity inhomogeneity, white matter underestimation, pial overestimation, and magnetic susceptibility artifact. Images were excluded for failure to pass all categories of QC, as well as for abnormal radiological findings (30).To account for outliers, the top and bottom 1% of each neuroimaging measure at each follow-up was winsorized.

### Covariates

Sex was a biological variable indicating a child’s sex assigned at birth. Only children assigned as male or female at birth were included for this analysis. Child ethnicity was categorized as Hispanic or non-Hispanic. Child race was indicated by the caregiver and categorized as White, Black, Asian, American Indian/Alaska Native or Native Hawaiian/Pacific Islander, More than one race, and Other. Child sex, race and ethnicity are time-invariant. Current parental education was modeled as a binary variable indicating whether the caregiver and/or their current partner had completed a bachelor’s degree. Current annual household income was categorized as less than $50,000, between $50,000 and $99,999, or at least $100,000. Parental education and annual household income are time-variant (referred to as current parental education and annual household income throughout).

### Analysis

Linear mixed effects models were conducted to examine relationships between GDM exposure (yes/no) and longitudinal changes in BMI and global brain measures (i.e., total cortical SA, mean CT, total cortical GMV, and subcortical GMV) as a function of individual maturation age (current age relative to onset of puberty). In models of BMI and global brain measures, the independent variables were GDM by maturation age interactions. For models with significant GDM X maturation age interactions, linear spline was computed where a notch was created at the age of onset of puberty for a GDM X spline interaction model. These models will be able to identify 1) group differences in slopes before puberty onset, 2) group differences in slopes after onset of puberty, and 3) group differences in change in slopes from pre-puberty to post-puberty.

In models of BMI, site ID, family ID, and subject ID were modeled as random effects. Fixed effect covariates included sex, race, ethnicity, current parental education, current family income, and current puberty. For models of structural MRI (sMRI) data, scanner ID, family ID, and subject ID were modeled as random effects. Fixed effect covariates included sex, race, ethnicity, current parental education, current family income, current puberty, and handedness. ICV was included as a covariate for volumetric models.

Mixed effects models were fitted in R (R version 4.0.3) (31) with the *lme4* package, and standardized betas were computed using the *parameters* package. *P*-values were calculated using the *lmerTest* package using Satterthwaite’s method, with correction for multiple comparisons using the Benjamini-Hochberg false discovery rate (FDR), at a corrected threshold for significance of *P*<0.05.

## Results

### Participants

After application of exclusions, the final analysis included 9,583 participants in either the BMI sample or the brain sample (details of sample sizes are included in **Supplementary** Figure 1). At baseline, 6.9% of participants were exposed to GDM *in utero*. **Table 1** shows demographic summaries of all participants included in either BMI or brain samples at baseline.

**Table 1.** Participant Baseline Characteristics.

|  | Un-exposed (N=8870) |  | GDM-Exposed (N=650) |  | All (N=9520)* |  | P** |
| --- | --- | --- | --- | --- | --- | --- | --- |
|  | N/Mean | %/SD | N/Mean | %/SD | N/Mean | %/SD |  |
| **Age (years)** | 9.9 | 0.6 | 9.9 | 0.6 | 9.9 | 0.6 | 0.574 |
| **Sex** |  |  |  |  |  |  | 0.814 |
| Male | 4,052 | 49.3% | 302 | 49.8% | 4,354 | 49.3% |  |
| Female | 4,175 | 50.7% | 304 | 50.2% | 4,479 | 50.7% |  |
| **Ethnicity** |  |  |  |  |  |  | 0.243 |
| Hispanic | 1,532 | 18.6% | 125 | 20.6% | 1,657 | 18.8% |  |
| Not Hispanic | 6,695 | 81.4% | 481 | 79.4% | 7,176 | 81.2% |  |
| **Race** |  |  |  |  |  |  | <0.001 |
| White | 5,636 | 68.5% | 371 | 61.2% | 6,007 | 68.0% |  |
| Black | 1,101 | 13.4% | 85 | 14.0% | 1,186 | 13.4% |  |
| Asian | 140 | 1.7% | 23 | 3.8% | 163 | 1.8% |  |
| AIANNHPI*** | 46 | 0.6% | 6 | 1.0% | 52 | 0.6% |  |
| More than one race | 995 | 12.1% | 90 | 14.9% | 1,085 | 12.3% |  |
| Other | 309 | 3.8% | 31 | 5.1% | 340 | 3.8% |  |
| **Family Income** |  |  |  |  |  |  | 0.023 |
| <\$50000 | 2,227 | 27.1% | 187 | 30.9% | 2,414 | 27.3% | |
| \$50000-\$99999 | 2,367 | 28.8% | 185 | 30.5% | 2,552 | 28.9% | |
| >=\$100000 | 3,633 | 44.2% | 234 | 38.6% | 3,867 | 43.8% | |
| **Parental Education** |  |  |  |  |  |  | 0.004 |
| Bachelor's Degree | 5,269 | 64.0% | 352 | 58.1% | 5,621 | 63.6% |  |
| No Bachelor's Degree | 2,958 | 36.0% | 254 | 41.9% | 3,212 | 36.4% |  |

\* Reported sample includes participants in either BMI analysis sample or brain sample at baseline.
\*\* Categorical variables by $\chi^2$ test, continuous variables by t-test. \*\*\*American Indian/Alaskan Native, Native Hawaiian/Pacific Islander.

### GDM exposure and BMI change

There was a significant exposure by maturation age interaction on BMI over time [β (95% CI) = 0.032 (0.008, 0.056), *P*=0.009]. BMI for un-exposed participants increased at a rate of 0.981 kg/m^2^ per year [95%CI = (0.956, 1.007)], whereas BMI for GDM-exposed participants increased at a rate of 1.063 kg/m^2^ per year [95%CI = (1.001, 1.125)] (**Table 2**, **Figure 1**).

**Table 2.**
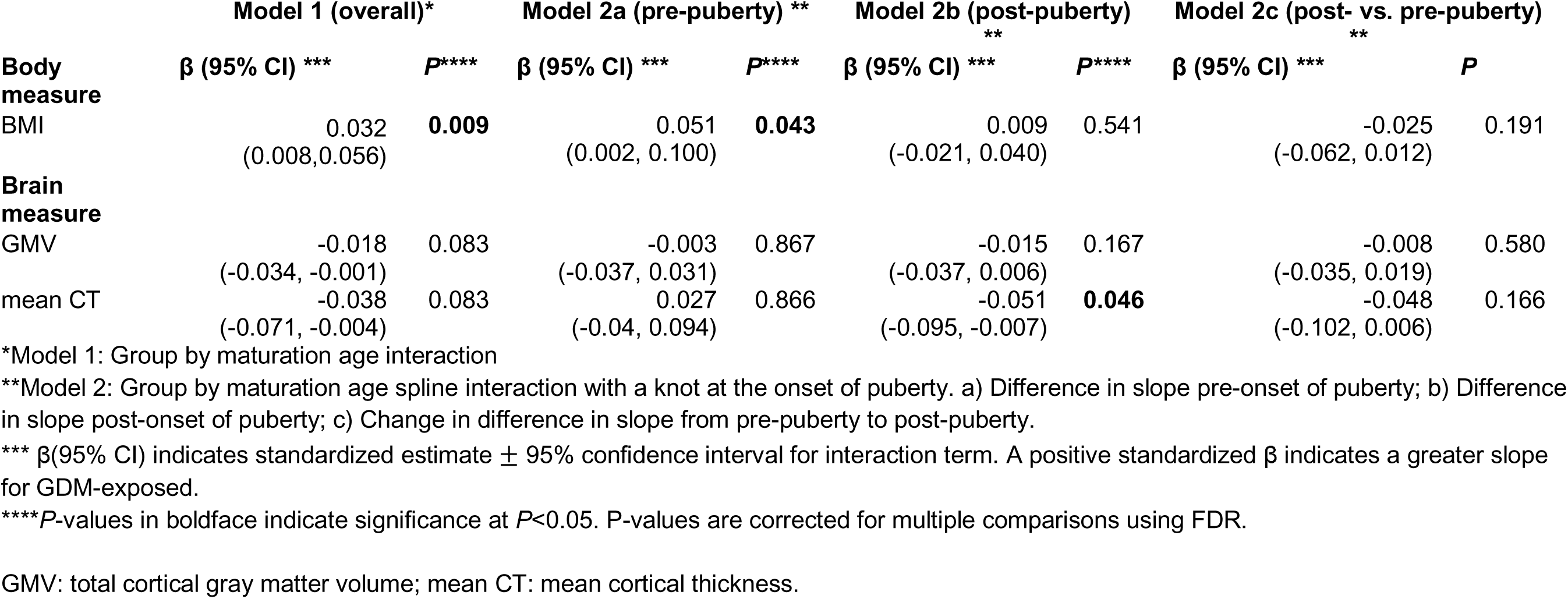
Relationships between gestational diabetes mellitus (GDM) exposure and BMI and global brain structural measures.

|  | Model 1 (overall)* |  | Model 2a (pre-puberty) ** |  | Model 2b (post-puberty) ** |  | Model 2c (post- vs. pre-puberty) ** |  |
| --- | --- | --- | --- | --- | --- | --- | --- | --- |
| **Body measure** | $\beta$ (95% CI) *** | *P* **** | $\beta$ (95% CI) *** | *P* **** | $\beta$ (95% CI) *** | *P* **** | $\beta$ (95% CI) *** | *P* |
| BMI | 0.032<br>(0.008,0.056) | **0.009** | 0.051<br>(0.002, 0.100) | **0.043** | 0.009<br>(-0.021, 0.040) | 0.541 | -0.025<br>(-0.062, 0.012) | 0.191 |
| **Brain measure** |  |  |  |  |  |  |  |  |
| GMV | -0.018<br>(-0.034, -0.001) | 0.083 | -0.003<br>(-0.037, 0.031) | 0.867 | -0.015<br>(-0.037, 0.006) | 0.167 | -0.008<br>(-0.035, 0.019) | 0.580 |
| mean CT | -0.038<br>(-0.071, -0.004) | 0.083 | 0.027<br>(-0.04, 0.094) | 0.866 | -0.051<br>(-0.095, -0.007) | **0.046** | -0.048<br>(-0.102, 0.006) | 0.166 |

\*Model 1: Group by maturation age interaction
\*\*Model 2: Group by maturation age spline interaction with a knot at the onset of puberty. a) Difference in slope pre-onset of puberty; b) Difference in slope post-onset of puberty; c) Change in difference in slope from pre-puberty to post-puberty.
\*\*\* $\beta$ (95% CI) indicates standardized estimate ± 95% confidence interval for interaction term. A positive standardized $\beta$ indicates a greater slope for GDM-exposed.
\*\*\*\**P*-values in boldface indicate significance at $P < 0.05$ . *P*-values are corrected for multiple comparisons using FDR.
GMV: total cortical gray matter volume; mean CT: mean cortical thickness.

There was an interaction of exposure by maturation age on change in BMI prior to onset of puberty [β(95%CI) = 0.051 (0.002, 0.100), *P* =0.043], but not after onset of puberty [β(95%CI) = 0.009 (−0.021, 0.040), *P* =0.541] according to a spline model (**Table 2**). Prior to puberty, BMI increased at an adjusted rate of 0.724 kg/m^2^ per year (95% CI = 0.687, 0.761) in un-exposed youth, compared with a rate of 0.854 kg/m^2^ per year (95% CI = 0.730, 0.977) among GDM-exposed participants. After the onset of puberty, BMI increased at an adjusted rate of 1.110 kg/m^2^ per year (95% CI = 1.081, 1.140) in un-exposed youth, compared with a rate of 1.134 kg/m^2^ per year (95% CI = 1.058, 1.211) among GDM-exposed participants.

### GDM exposure and sMRI change

There were uncorrected significant interactions of exposure by maturation age for total GMV [β (95% CI) = −0.018 (−0.034, −0.001), uncorrected *P*=0.042] and mean CT [β (95% CI) = −0.038 (−0.071, −0.004), uncorrected *P*=0.027]. Interactions were not significant for total subcortical GMV [β (95% CI) = −0.001 (−0.015, 0.013), *P*=0.906] and SA [β (95% CI) =0.004 (−0.007, 0.016), *P*=0.633].

According to spline models, there was a significant exposure by maturation age interaction in mean CT over time after onset of puberty [β (95% CI) = −0.051 (−0.095, −0.007), *P*=0.046)] but not before [β (95% CI) = 0.027 (−0.040, −0.094), *P*=0.433)]. Prior to puberty, mean CT changed at an adjusted rate of −0.012 mm per year (95% CI = −0.013, −0.011) in un-exposed participants, and at a rate of −0.011 mm per year (95% CI = −0.013, −0.008) in GDM-exposed participants; whereas after the onset of puberty, mean CT changed at a rate of −0.020 mm per year (95% CI = −0.021, −0.019) in un-exposed participants, and at a rate of −0.022 mm per year (95% CI = −0.024, −0.020) in GDM-exposed participants (**Figure 2**). There was a trend of significance in the change in slope between groups from before to after onset of puberty in mean CT [ β (95% CI) = −0.048 (−0.102, 0.006), *P*=0.083)]. There were no significant group by maturation age interactions in total GMV over time both before [β (95% CI) = −0.003 (−0.037, 0.031), *P*=0.867] and after onset of puberty [β (95% CI) = −0.015 (−0.037, 0.006), *P*=0.167]. The change in slope between groups from before to after onset of puberty was not significant in total GMV [β (95% CI) = −0.008 (−0.035, 0.019), *P*=0.580].

**Figure 2.**
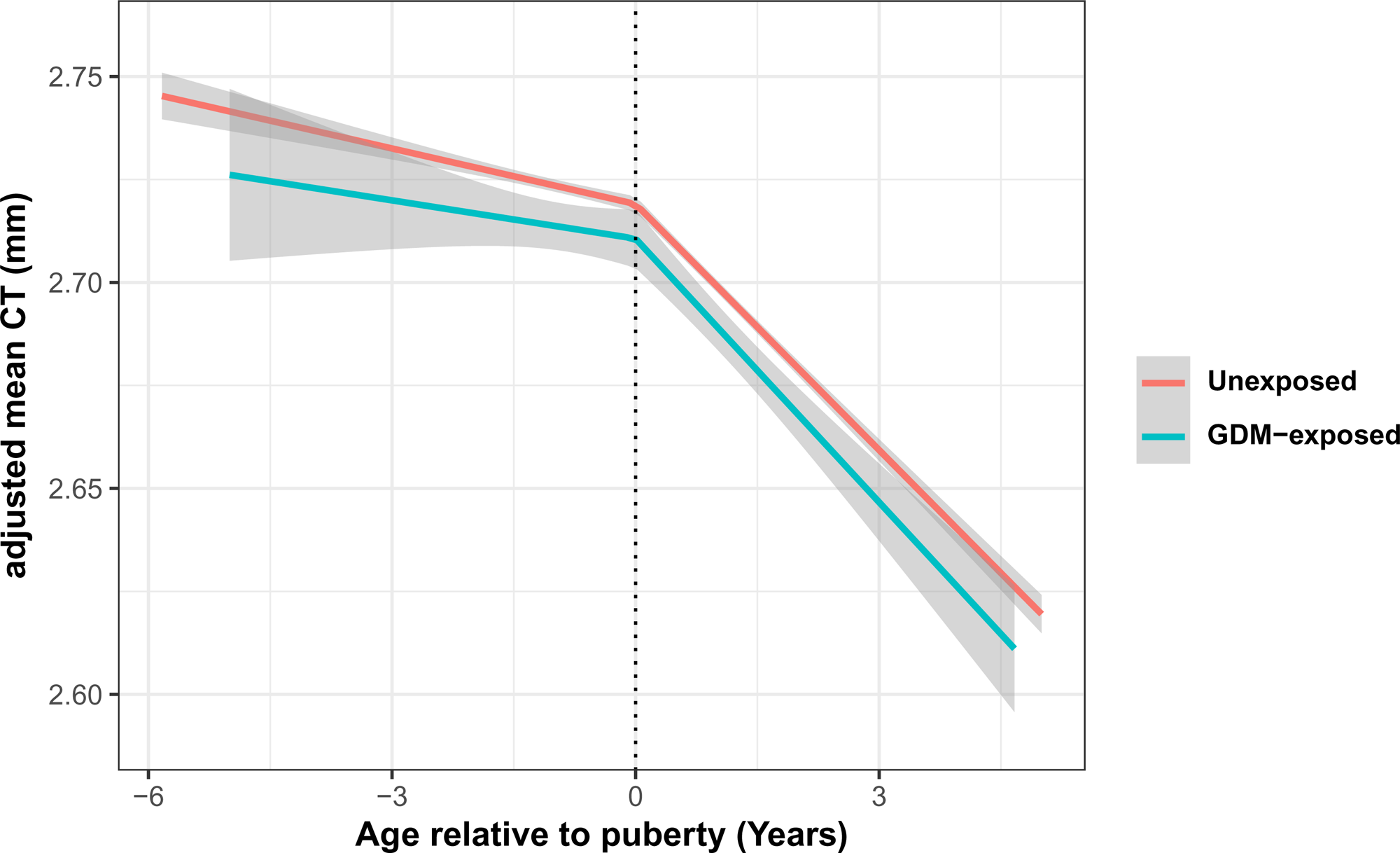
Relationship of maturation age (age relative to onset of puberty, x-axis) by gestational diabetes mellitus (GDM) exposure status interaction in mean cortical thickness (CT) with adjustment for family ID and scanner ID as random effects, and handedness, sex, race, ethnicity, current pubertal status, current family income, and current parental education as fixed effects. Lines represent linear spline mixed-effects regression fit with a knot at the age of onset of puberty, and shaded region represents one standard error (SE) region of regression fit.

## Discussion

This is the first multi-site study to investigate longitudinal associations between prenatal exposure to GDM, BMI, and brain structure, in a diverse sample of youth over a period of late childhood into mid-adolescence. We found that prenatal exposure to GDM was associated with greater increases in BMI between 9 and 15 years of age, which was driven by larger BMI growth among GDM-exposed offspring prior to, but not after, onset of puberty. GDM exposure was associated with greater decreases in mean CT over the period of 9-15 years of age. Furthermore, groups differed in CT following the onset of puberty (but not before), with a faster rate of declining among those prenatally exposed to GDM compared to the un-exposed offspring. These results suggest that prenatal exposure to GDM was associated with greater pre-pubertal increases in BMI and faster post-pubertal decreases in cortical thickness during peri-adolescence.

Consistent with prior longitudinal studies (13,32), we found that prenatal GDM exposure was associated with greater increases in BMI over time. Previous longitudinal studies have found greater increases in BMI among GDM-exposed offspring (vs. un-exposed) from birth to 15 years of age, with differential trajectories appearing largest in adolescence. For example, a longitudinal study showed that offspring exposed to GDM had increases in BMI beginning around 10 years of age (33), compared to a reference population without GDM exposure, and another study reported that divergence in BMI growth trajectory between groups was the greatest between 10 and 13 years (16,17).

The time window described by prior studies includes the beginning of hormonal and bodily changes associated with the onset of puberty. In the current study, we found greater BMI growth in GDM-exposed offspring (vs. un-exposed) from late-childhood into mid-adolescence (9-15 years), and these increases were observed *prior to* onset of puberty only. Taken together with previous work, these results suggest that GDM exposure is associated with greater increases in BMI during peri-adolescence, and specific time window when groups diverge in BMI trajectory may occur around middle childhood before onset of puberty.

Prior cross-sectional studies have reported that prenatal GDM exposure was associated with smaller global and regional brain structural correlates (34). To our best knowledge, this is the first longitudinal study investigating relationships between GDM exposure and trajectory of structural brain indices. Brain structure measures such as CT, GMV and SA peak between 1 and 9 years of age (18,35). Between the ages of 9 and 15, it is expected that these measures would decline. Here, we found that GDM exposure was associated with faster declines in total GMV and mean CT between ages of 9 and 15, suggesting a deviation from normal brain development trajectory in GDM-exposed offspring. Furthermore, we found that group differences in mean CT over time was driven by changes beginning *after* onset of puberty but not before, with GDM-exposed participants exhibiting a faster declining rate than un-exposed offspring. One possible explanation for accelerated post-pubertal changes in brain structure development is a “double-hit” or “two-hit” hypothesis, which has been hypothesized as an explanation for adolescent-onset mental disorders (36–38). According to this theory, the interaction of multiple adversities, beginning with perinatal stressors, and followed later by peri-pubertal stressors, trigger adolescent onset of negative health outcomes. Intrauterine exposure to GDM has been shown to impact the hypothalamic-pituitary-adrenal (HPA) axis negatively, a neuroendocrine system instrumental in pubertal development. Perinatal alterations to the HPA axis may then be related to accelerated neurological and physical development in association with puberty (17), the second “hit.” These data support promotion of interventions in late childhood, prior to onset of puberty, to address risk of altered brain development in offspring with prenatal GDM exposure.

We recognized several limitations of this study. GDM-exposure and pubertal stage of development were assessed based on parental self-reports, which could be susceptible to recall biases. Maternal characteristics in pregnancy such as severity and treatment of diabetes are not available in the ABCD dataset, thus we are not able to assess their modulating effects on BMI and brain structure development in youth with and without GDM exposure. Covariates included in this study are limited to availability in the ABCD study. While we focused on global brain structural measures, future study is warranted to study other regional imaging markers in relation to prenatal exposure to GDM. Lastly, we did not investigate a potential mediating role of brain in the longitudinal associations of GDM exposure with BMI.

## Conclusion

In this large multi-site study including over 9,500 youth, we observed that prenatal exposure to GDM is associated with greater increases in BMI and faster declines in cortical thickness between the ages of 9-15. Faster BMI growth is observed prior to onset of puberty, while faster cortical thinning is found after the onset of puberty. Our data suggest that puberty may serve as a trigger for altered adolescent brain development in offspring prenatally exposed to GDM.

## Author Contributions

S.L. and E.H. performed the statistical analysis and drafted the manuscript. S.L., E.H., and T.P. provided review, commentary, and revisions to the manuscript, approved the final manuscript as submitted. S.L. is the guarantor of this work and, as such, had full access to all the data in the study and takes responsibility for the integrity of the data and the accuracy of the data analysis.

## Data Availability

All data produced in the present study are available upon reasonable request to the authors

## Acknowledgments

The authors acknowledge and give thanks to the participants of the ABCD Study and their families.

Research described in this study was supported by the National Institutes of Health National Institute of Diabetes and Digestive and Kidney Diseases R01DK137899 (PI: SL), K01DK115638 (PI: SL), R03DK129186 (PI: SL). Data used in the preparation of this article were obtained from the Adolescent Brain Cognitive Development® (ABCD) Study (https://abcdstudy.org), held in the NIMH Data Archive (NDA). This is a multisite, longitudinal study designed to recruit more than 10,000 children age 9-10 and follow them over 10 years into early adulthood. The ABCD Study® is supported by the National Institutes of Health and additional federal partners under award numbers U01DA041048, U01DA050989, U01DA051016, U01DA041022, U01DA051018, U01DA051037, U01DA050987, U01DA041174, U01DA041106, U01DA041117, U01DA041028, U01DA041134, U01DA050988, U01DA051039, U01DA041156, U01DA041025, U01DA041120, U01DA051038, U01DA041148, U01DA041093, U01DA041089, U24DA041123, U24DA041147. A full list of supporters is available at https://abcdstudy.org/federal-partners.html. A listing of participating sites and a complete listing of the study investigators can be found at https://abcdstudy.org/consortium_members/. ABCD consortium investigators designed and implemented the study and/or provided data but did not necessarily participate in the analysis or writing of this report. This manuscript reflects the views of the authors and may not reflect the opinions or views of the NIH or ABCD consortium investigators. The ABCD data repository grows and changes over time. The ABCD data used in this report came from http://dx.doi.org/10.15154/z563-zd24.

