## Supplementary Figure for "Prenatal Exposure to Gestational Diabetes Mellitus is Associated with Greater Pre-pubertal BMI Growth and Faster Post-pubertal Cortical Thinning During Peri-adolescence"

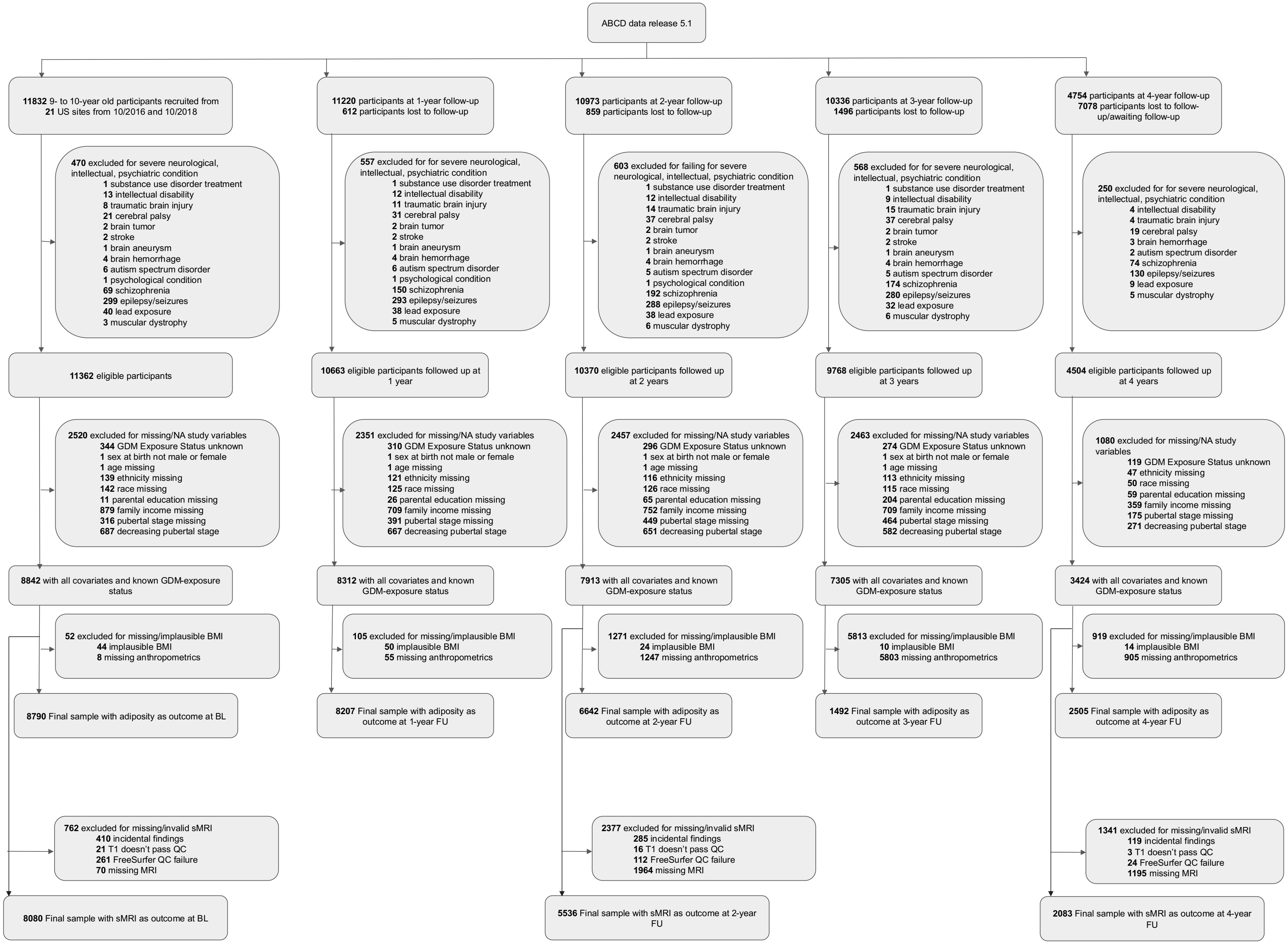

ABCD data release 5.1
11832 9- to 10-year old participants recruited from 21 US sites from 10/2016 and 10/2018
11220 participants at 1-year follow-up 612 participants lost to follow-up
10973 participants at 2-year follow-up 859 participants lost to follow-up
10336 participants at 3-year follow-up 1496 participants lost to follow-up
4754 participants at 4-year follow-up 7078 participants lost to follow-up/awaiting follow-up
470 excluded for severe neurological, intellectual, psychiatric condition
1 substance use disorder treatment
13 intellectual disability
8 traumatic brain injury
21 cerebral palsy
2 brain tumor
2 stroke
1 brain aneurysm
4 brain hemorrhage
6 autism spectrum disorder
1 psychological condition
69 schizophrenia
299 epilepsy/seizures
40 lead exposure
3 muscular dystrophy
557 excluded for for severe neurological, intellectual, psychiatric condition
1 substance use disorder treatment
12 intellectual disability
11 traumatic brain injury
31 cerebral palsy
2 brain tumor
2 stroke
1 brain aneurysm
4 brain hemorrhage
6 autism spectrum disorder
1 psychological condition
150 schizophrenia
293 epilepsy/seizures
38 lead exposure
5 muscular dystrophy
603 excluded for failing for severe neurological, intellectual, psychiatric condition
1 substance use disorder treatment
12 intellectual disability
14 traumatic brain injury
37 cerebral palsy
2 brain tumor
2 stroke
1 brain aneurysm
4 brain hemorrhage
5 autism spectrum disorder
1 psychological condition
192 schizophrenia
288 epilepsy/seizures
38 lead exposure
6 muscular dystrophy
568 excluded for for severe neurological, intellectual, psychiatric condition
1 substance use disorder treatment
9 intellectual disability
15 traumatic brain injury
37 cerebral palsy
2 brain tumor
2 stroke
1 brain aneurysm
4 brain hemorrhage
5 autism spectrum disorder
174 schizophrenia
280 epilepsy/seizures
32 lead exposure
6 muscular dystrophy
250 excluded for for severe neurological, intellectual, psychiatric condition
4 intellectual disability
4 traumatic brain injury
19 cerebral palsy
3 brain hemorrhage
2 autism spectrum disorder
74 schizophrenia
130 epilepsy/seizures
9 lead exposure
5 muscular dystrophy
11362 eligible participants
10663 eligible participants followed up at 1 year
10370 eligible participants followed up at 2 years
9768 eligible participants followed up at 3 years
4504 eligible participants followed up at 4 years
2520 excluded for missing/NA study variables
344 GDM Exposure Status unknown
1 sex at birth not male or female
1 age missing
139 ethnicity missing
142 race missing
11 parental education missing
879 family income missing
316 pubertal stage missing
687 decreasing pubertal stage
2351 excluded for missing/NA study variables
310 GDM Exposure Status unknown
1 sex at birth not male or female
1 age missing
121 ethnicity missing
125 race missing
26 parental education missing
709 family income missing
391 pubertal stage missing
667 decreasing pubertal stage
2457 excluded for missing/NA study variables
296 GDM Exposure Status unknown
1 sex at birth not male or female
1 age missing
116 ethnicity missing
126 race missing
65 parental education missing
752 family income missing
449 pubertal stage missing
651 decreasing pubertal stage
2463 excluded for missing/NA study variables
274 GDM Exposure Status unknown
1 sex at birth not male or female
1 age missing
113 ethnicity missing
115 race missing
204 parental education missing
709 family income missing
464 pubertal stage missing
582 decreasing pubertal stage
1080 excluded for missing/NA study variables
119 GDM Exposure Status unknown
47 ethnicity missing
50 race missing
59 parental education missing
359 family income missing
175 pubertal stage missing
271 decreasing pubertal stage
8842 with all covariates and known GDM-exposure status
8312 with all covariates and known GDM-exposure status
7913 with all covariates and known GDM-exposure status
7305 with all covariates and known GDM-exposure status
3424 with all covariates and known GDM-exposure status
52 excluded for missing/implausible BMI
44 implausible BMI
8 missing anthropometrics
105 excluded for missing/implausible BMI
50 implausible BMI
55 missing anthropometrics
1271 excluded for missing/implausible BMI
24 implausible BMI
1247 missing anthropometrics
5813 excluded for missing/implausible BMI
10 implausible BMI
5803 missing anthropometrics
919 excluded for missing/implausible BMI
14 implausible BMI
905 missing anthropometrics
8790 Final sample with adiposity as outcome at BL
8207 Final sample with adiposity as outcome at 1-year FU
6642 Final sample with adiposity as outcome at 2-year FU
1492 Final sample with adiposity as outcome at 3-year FU
2505 Final sample with adiposity as outcome at 4-year FU
762 excluded for missing/invalid sMRI
410 incidental findings
21 T1 doesn't pass QC
261 FreeSurfer QC failure
70 missing MRI
2377 excluded for missing/invalid sMRI
285 incidental findings
16 T1 doesn't pass QC
112 FreeSurfer QC failure
1964 missing MRI
1341 excluded for missing/invalid sMRI
119 incidental findings
3 T1 doesn't pass QC
24 FreeSurfer QC failure
1195 missing MRI
8080 Final sample with sMRI as outcome at BL
5536 Final sample with sMRI as outcome at 2-year FU
2083 Final sample with sMRI as outcome at 4-year FU

**Figure S1.** Sample Exclusion Flow Chart. Flowchart depicts exclusions of ineligible participants based on *four* categories of criteria: 1) medical history, including *initial screening, 2*) missing data or unknown status for study variables including status of Gestational Diabetes Mellitus (GDM) exposure, age at interview, sex at birth, race, ethnicity, family income, parental education history, and pubertal stage assessment, 3) body mass index (BMI) out of range, and quality control (QC) for structural brain (T1) image.
